# Association Between ACEIs or ARBs Use and Clinical Outcomes in COVID-19 Patients: A Systematic Review and Meta-analysis

**DOI:** 10.1101/2020.06.03.20120261

**Authors:** Carlos Diaz-Arocutipa, Jose Saucedo-Chinchay, Adrian V. Hernandez

## Abstract

**Importance:** There is a controversy regarding whether or not to continue angiotensin-converting enzyme inhibitors (ACEIs) and angiotensin receptor blockers (ARBs) in patients with coronavirus disease 2019 (COVID-19).

**Objective:** To evaluate the association between ACEIs or ARBs use and clinical outcomes in COVID-19 patients.

**Data Sources:** Systematic search of the PubMed, Embase, Scopus, Web of Science, and Cochrane Central Register of Controlled Trials from database inception to May 31, 2020. We also searched the preprint servers medRxiv and SSNR for additional studies.

**Study Selection:** Observational studies and randomized controlled trials reporting the effect of ACEIs or ARBs use on clinical outcomes of adult patients with COVID-19.

**Data Extraction and Synthesis:** Risk of bias of observational studies were evaluated using the Newcastle-Ottawa Scale. Meta-analyses were performed using a random-effects models and effects expressed as Odds ratios (OR) and mean differences with their 95% confidence interval (95%CI). If available, adjusted effects were pooled.

**Main Outcomes and Measures:** The primary outcome was all-cause mortality and secondary outcomes were COVID-19 severity, hospital discharge, hospitalization, intensive care unit admission, mechanical ventilation, length of hospital stay, and troponin, creatinine, procalcitonin, C-reactive protein (CRP), interleukin-6 (IL-6), and D-dimer levels.

**Results:** 40 studies (21 cross-sectional, two case-control, and 17 cohorts) involving 50615 patients were included. ACEIs or ARBs use was not associated with all-cause mortality overall (OR 1.11, 95%CI 0.77-1.60, p=0.56), in subgroups by study design and using adjusted effects. ACEI or ARB use was independently associated with lower COVID-19 severity (aOR 0.56, 95%CI 0.37-0.87, p<0.01). No significant associations were found between ACEIs or ARBs use and hospital discharge, hospitalization, mechanical ventilation, length of hospital stay, and biomarkers.

**Conclusions and Relevance:** ACEIs or ARBs use was not associated with higher all-cause mortality in COVID-19. However, ACEI or ARB use was independently associated with lower COVID-19 severity. Our results support the current international guidelines to continue the use of ACEIs and ARBs in COVID-19 patients with hypertension.

**Key points:** *Question:* What is the association between angiotensin-converting enzyme inhibitors (ACEIs) or angiotensin receptor blockers (ARBs) use and clinical outcomes in coronavirus disease 2019 (COVID-19) patients?

*Findings:* In this systematic review and meta-analysis of 40 observational studies, the use of ACEIs or ARBs was not associated with higher all-cause mortality in COVID-19 patients. Additionally, ACEIs or ARBs use was independently associated with lower COVID-19 severity.

*Meaning:* These results support the current international guidelines to continue the use of ACEIs and ARBs in COVID-19 patients with hypertension.

## Introduction

Coronavirus disease 2019 (COVID-19) is a global pandemic involving more than 185 countries.^1^ This disease is caused by the severe acute respiratory coronavirus-2 (SARS-CoV-2) and was first detected in Wuhan, China in December, 2019.^2^ The infection by SARS-CoV-2 is caused by the binding of the viral spike glycoprotein to the angiotensin-converting enzyme 2 (ACE2).^3^ In humans, ACE2 is ubiquitously expressed with predominance in the lungs, heart, kidneys, and vascular system.^4^ ACE2 is a major component of the renin-angiotensin system (RAS) that acts as a carboxypeptidase converting angiotensin II (Ang II) into angiotensin 1-7 (Ang 1-7).^5^ The degradation of Ang II by ACE2 regulates negatively the RAS activation and attenuates the vasoconstrictive, pro-oxidant, pro-fibrotic, and pro-inflammatory actions mediated by Ang II.^6^ The RAS is considered a complex system that requires a balanced interplay between two counter-regulatory axes (ACE2/Ang 1-7/MasR and ACE/Ang II/AT_1_R).^6^ Moreover, there is evidence that ACE2 has a protective physiological role in many organs, including lungs and heart, and its imbalance can be lead to disease states.^5^

Angiotensin-converting enzyme inhibitors (ACEIs) and angiotensin receptor blockers (ARBs) are widely used in clinical practice for the treatment of hypertension, heart failure, and diabetic nephropathy.^7^ It has been hypothesized that these drugs could increase the risk of infection and severity in COVID-19 patients.^8-10^ However, major international cardiology societies have recommended not to discontinue the use of ACEIs and ARBs in COVID-19 patients with hypertension due to a lack of clinical evidence.^11^

Recently, several studies that evaluated the effect of RAS inhibitors on COVID-19 have been published. Therefore, we performed a systematic review and meta-analysis to evaluate the association between ACEIs or ARBs use and clinical outcomes in COVID-19 patients.

## Methods

This review was reported according to the MOOSE (Meta-analysis of Observational Studies in Epidemiology) guidelines (eTable 1)^12^ and was registered in PROSPERO database (CRD42020177848).

### Search strategy

We searched PubMed, Embase, Scopus, Web of Science, and Cochrane Central Register of Controlled Trials. The preprint servers medRxiv and SSNR were also searched. The search was conducted from inception to April 4, 2020, and updated on May 31, 2020. The complete search strategy is available in eTable 2. There were no restrictions on language. We conducted hand searches of reference lists of included studies and relevant reviews articles to identify further eligible studies. Additionally, clinicaltrials.gov registry was searched for finished as well as ongoing randomized controlled trials (RCTs).

### Eligibility criteria

We included observational studies and RCTs that evaluated the association between ACEIs or ARBs use and at least one clinical outcome in COVID-19 patients (≥18 years) diagnosed by reverse transcription-polymerase chain reaction. Case reports, case series, systematic reviews, narrative reviews, commentaries, and abstracts were excluded.

### Study Selection

Two authors (CDA and JSC) downloaded all articles from electronic search to EndNote X8 and duplicates were removed. Titles and abstracts were independently screened by two authors (CDA and JSC) to identify potentially relevant studies. Two authors (CDA and JSC) independently screened the full-text and registered reasons for the exclusion. Any disagreement was resolved by consensus.

### Outcomes

The primary outcome was all-cause mortality and the secondary outcomes were COVID-19 severity, hospital discharge, hospitalization, intensive care unit (ICU) admission, mechanical ventilation, length of hospital stay, troponin, creatinine, procalcitonin, C-reactive protein (CRP), interleukin-6 (IL-6) and D-dimer. We used author-reported definitions for all outcomes.

### Data Extraction

Information from each study was independently extracted by two authors (CDA and JSC) using a standardized data extraction form and any disagreement was resolved by consensus. If additional data was needed, we contacted the corresponding author through email. We extracted the following data: author, publication year, country, study design, sample size, eligibility criteria, age, sex, comorbidities, ACEIs or ARBs use, and primary and secondary outcomes. If available, unadjusted and adjusted effect measures were also extracted.

### Risk of bias assessment

The Newcastle-Ottawa Scale (NOS) was used to assess the risk of bias in case-control and cohort studies.^13^ Each study was classified in the following groups: low risk of bias (8-9 points), moderate risk of bias (5-7 points), and high risk of bias (0-4 points). For cross-sectional studies, we used an adapted version of NOS^14^ and each study was assigned in the following groups: low risk of bias (8-10 points), moderate risk of bias (5-7 points), and high risk of bias (0-4 points). The risk of bias was independently assessed by two authors (CDA and JSC) and any disagreement was resolved by consensus.

### Statistical analysis

We performed all meta-analyses using random-effects models. Between-study variance was estimated using the Paule-Mandel estimator.^15^ We pooled odds ratios (OR) and mean differences (MD) with their 95% confidence intervals (95%CIs) for binary and continuous outcomes, respectively. In case of studies have only reported median and interquartile range, then mean and standard deviation were estimated using the method published by Wan et al.^16^ As exploratory analyses, we combined adjusted effects from studies that included a minimum set of confounding variables (age, sex, and cardiovascular comorbidities) in their multivariate models. Heterogeneity among studies was evaluated using the chi-squared test (threshold p<0.10) and I^2^ statistic. Heterogeneity was defined as low if I^2^<30%, moderate if I^2^=30-60%, and high if I^2^>60%. Funnel plots were used to evaluate publication bias and the Egger’s test was performed to measure asymmetry of funnel plots only if 10 or more studies were included.^17^ Subgroup analyses were conducted according to study design (cross-sectional vs cohort). In post hoc sensitivity analyses, we adjusted all 95%CIs using the Hartung-Knapp method to address possible type I error with the conventional random-effects approach.^18^ All meta-analyses were conducted using the meta package from R 3.6.3. A two-tailed p<0.05 was considered as statistically significant.

## Results

### Study selection

Our search strategy identified initially 110 articles. After removal of duplicates, 87 articles remained. After screening of studies by title/abstract, 36 articles were excluded. After full-text revision of 51 articles, 11 articles were excluded. A total of 40 studies were selected for analysis (21 cross-sectional, two case-control, and 17 cohort studies) (Figure 1). Only one contacted author provided additional information on mortality.^19^

**Figure 1.**
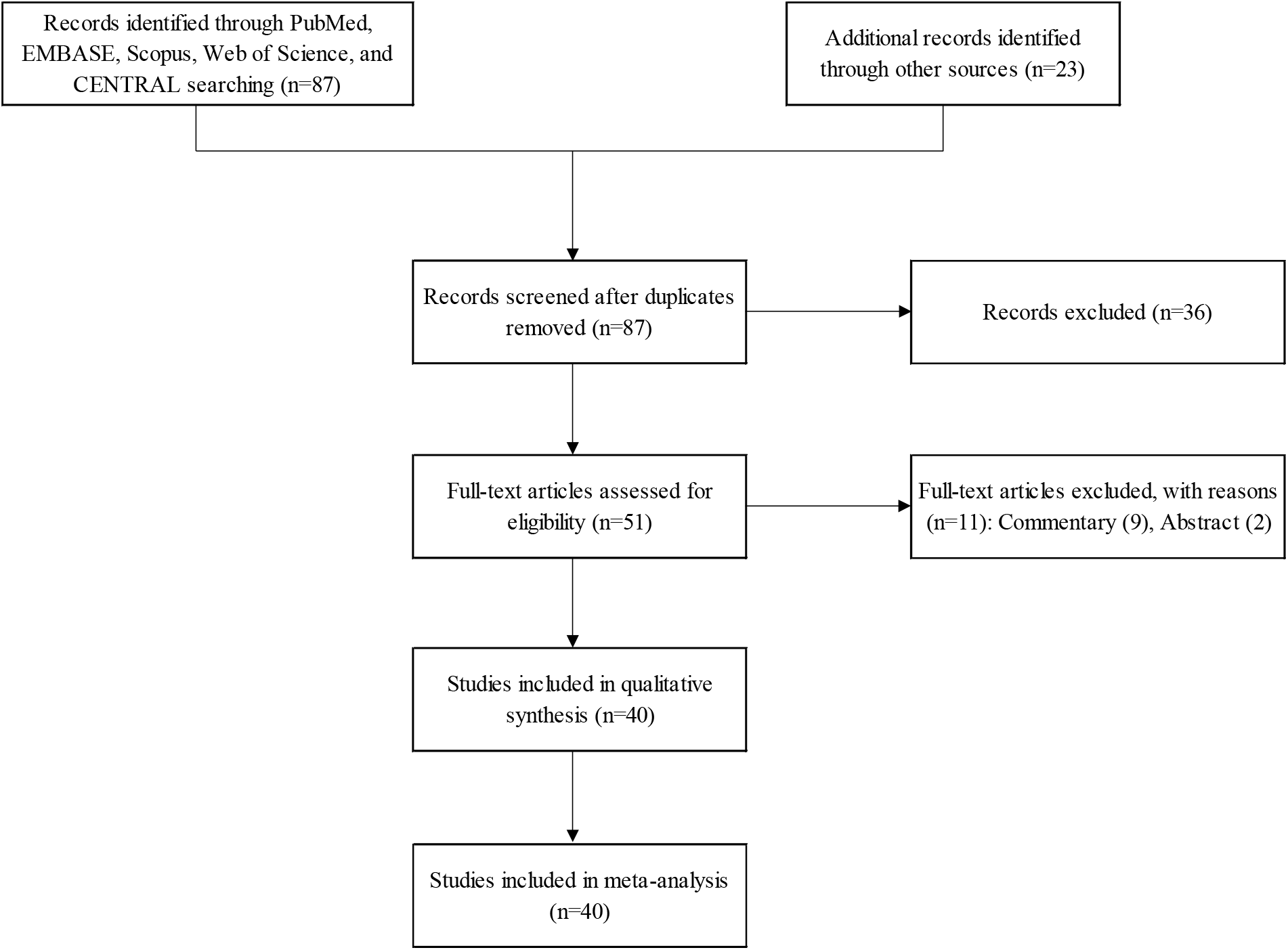
Flow diagram of study selection.

### Study Characteristics

Main characteristics of the 40 included studies (n=50615) were summarized in Table 1. The exposure variable in almost all studies was the chronic use of ACEIs and ARBs (i.e. before hospital admission) as registered in medical records, although in two studies^20,21^ it was defined as in-hospital use. Also, only six studies^22-27^ reported that ACEIs and ARBs were not discontinued during hospitalization. The definition of outcomes was the same in almost all included studies. In contrast, the definition of COVID-19 severity was very heterogeneous across the studies due to different clinical guidelines used for management of COVID-19 in each country. The most common criteria for COVID-19 severity was critical/severe vs mild/moderate which was used in five studies^24,28-31^ (Table 1).

**Table 1.** Characteristics of included studies.

| Study | Country | Study design | Sample size | Eligibility criteria | Age, years | Male | Comorbidities | ACEIs or ARBs use | Outcomes | Definition of mortality | Definition of COVID-19 severity |
| --- | --- | --- | --- | --- | --- | --- | --- | --- | --- | --- | --- |
| Guo et al,<sup>25</sup> 2020 | China | Cross-sectional | 187 | Patients with COVID-19 and who were treated and discharged or died during hospitalization | 58.5 ± 14.6<sup>b</sup> | 49% | Hypertension (33%), diabetes (15%), CAD (11.2%) | 10% | All-cause mortality | In-hospital death | NR |
| Peng et al,<sup>73</sup> 2020 | China | Cross-sectional | 112 | Patients with COVID-19 and cardiovascular diseases | 62 (55 - 67)<sup>c</sup> | 47% | Hypertension (82%), diabetes (20%), CAD (55%), HF (36%) | 20% | All-cause mortality, COVID-19 severity | In-hospital death | Critical vs mild/severe |
| Meng et al,<sup>27</sup> 2020 | China | Cross-sectional | 42 | Patients with COVID-19 and hypertension | 64.5 (55.8-69)<sup>c</sup> | 57% | Hypertension (100%), diabetes (14%), CAD (19%) | 40% | All-cause mortality, COVID-19 severity, troponin, creatinine, procalcitonin, IL-6, D-dimer | In-hospital death | Severe vs moderate |
| Liu et al,<sup>74</sup> 2020 | China | Cross-sectional | 78 | Adult patients (≥18 years) with COVID-19 and hypertension | 65.2 ± 10.7<sup>b</sup> | 55% | Hypertension (100%) | 28% | COVID-19 severity | NR | Severe vs mild |
| Zeng et al,<sup>39</sup> 2020 | China | Cross-sectional | 75<sup>a</sup> | Patients with COVID-19 | 67 ± 11<sup>b</sup> | 47% | Hypertension (100%), diabetes (31%), CVD (21%) | 37% | All-cause mortality, COVID-19 severity, hospitalization, hospital discharge, length of hospital stay | In-hospital death | Severe vs non-severe |
| Yun et al,<sup>75</sup> 2020 | China | Cross-sectional | 113<sup>a</sup> | Patients with COVID-19 | 53 (40-64)<sup>c</sup> | 57% | Hypertension (100%) | 29% | COVID-19 severity | NR | Critical/severe vs moderate |
| Li et al,<sup>26</sup> 2020 | China | Cross-sectional | 362 | Patients with COVID-19 and hypertension | 66 (59-73)<sup>c</sup> | 52% | Hypertension (100%), diabetes (35%), CAD (17%), HF (3%) | 32% | All-cause mortality, COVID-19 severity, length of hospital stay, troponin, creatinine, procalcitonin, CRP, IL-6, D-dimer | In-hospital death | Severe vs non-severe |
| Argenziano et al,<sup>41</sup> 2020 | USA | Cross-sectional | 1000 | Consecutive patients with COVID-19 who received emergency or inpatient care | 61.7 ± 17.5<sup>b</sup> | 60% | Hypertension (60%), diabetes (37%), CAD (13%), HF (10%) | 28% | ICU admission | NR | NR |
| Chen et al,<sup>20</sup> 2020 | China | Cross-sectional | 123 | Patients with COVID-19 | 57.7 ± 12.7<sup>b</sup> | 43% | Hypertension (33%), diabetes (11%), CAD (12%) | 9% | All-cause mortality | In-hospital death | NR |
| Ashraf et al,<sup>22</sup> 2020 | Iran | Cross-sectional | 100 | Hospitalized patients with COVID-19 | 58 (48-68)<sup>c</sup> | 64% | Hypertension (26%), diabetes (26%), CAD (19%) | 19% | COVID-19 severity | NR | Critical vs non-critical |
| Richardson et al,<sup>44</sup> 2020 | USA | Cross-sectional | 1366<sup>a</sup> | Hospitalized patients with COVID-19 | 63 (52-75)<sup>c</sup> | 60% | Hypertension (100%) | 30% | All-cause mortality, ICU admission, mechanical ventilation | In-hospital death | NR |
| Ip et al,<sup>76</sup> 2020 | USA | Cross-sectional | 1129<sup>a</sup> | Hospitalized patients with COVID-19 and hypertension | NR | NR | Hypertension (100%) | 41% | All-cause mortality | In-hospital death | NR |
| Tedeschi et al,<sup>36</sup> 2020 | Italy | Cross-sectional | 311<sup>a</sup> | Hospitalized patients with COVID-19 and hypertension | 76 (67-83)<sup>c</sup> | 72% | Hypertension (100%), diabetes (24%), CVD (42%) | 56% | All-cause mortality | In-hospital death | NR |
| Wang et al,<sup>77</sup> 2020 | China | Cross-sectional | 344 | Patients admitted to ICU with COVID-19 | 64 (52-72)<sup>c</sup> | 52% | Hypertension (41%), diabetes (19%), CVD (12%) | 18% | All-cause mortality | In-hospital death | NR |
| Reynolds et al,<sup>78</sup> 2020 | USA | Cross-sectional | 2573<sup>a</sup> | Patients with COVID-19 | 64 (54-75)<sup>c</sup> | 52% | Hypertension (100%) | 50% | COVID-19 severity | NR | ICU admission, mechanical ventilation, or death vs none |
| Mancia et al,<sup>79</sup> 2020 | Italy | Cross-sectional | 6272<sup>a</sup> | Patients with COVID-19 | 68 ± 13<sup>b</sup> | 63% | Hypertension (58%), CAD (8%), HF (5%) | 46% | COVID-19 severity | NR | Critical/fatal vs mild/mode rate |
| Jurado et al,<sup>80</sup> 2020 | Spain | Cross-sectional | 290<sup>a</sup> | Hospitalized patients with COVID-19 | NR | NR | Hypertension (100%) | 67% | COVID-19 severity | NR | Severe vs mild/mode rate |
| Regina et al,<sup>45</sup> 2020 | Switzerland | Cross-sectional | 200 | Hospitalized adult patients (≥18 years) with COVID-19 | 70 (55-81)<sup>c</sup> | 60% | Hypertension (44%), diabetes (22%), CAD (18%), CKD (14%) | 26% | Mechanical ventilation | NR | NR |
| Tan et al,<sup>29</sup> 2020 | China | Cross-sectional | 100 | Hospitalized patients with COVID-19 and hypertension | NR | 51% | Hypertension (100%), diabetes (28%), CAD (18%), CKD (9%) | 31% | All-cause mortality, COVID-19 severity, hospital discharge, mechanical ventilation, length of hospital stay, CRP, D-dimer | In-hospital death | Critical/severe vs mild/mode rate |
| Zhou et al,<sup>46</sup> 2020 | China | Cross-sectional | 36<sup>a</sup> | Hospitalized patients with COVID-19 and hypertension | 64.8 ± 10.1<sup>b</sup> | 53% | Hypertension (100%) | 42% | All-cause mortality, length of hospital stay | In-hospital death | NR |
| Conversano et al,<sup>81</sup> 2020 | Italy | Cross-sectional | 191 | Hospitalized adult patients with COVID-19 pneumonia | 63.4 ± 14.9<sup>b</sup> | 69% | Hypertension (50%), diabetes (15%), CAD (15%), HF (5%), CKD (26%) | 36% | All-cause mortality | In-hospital death | NR |
| Yan et al,<sup>30</sup> 2020 | China | Case-control | 610 | Consecutive adult patients with COVID-19 | 48.7 ± 14.2<sup>b</sup> | 51% | Hypertension (22%), diabetes (10%), CVD (3%) | 42% | COVID-19 severity | NR | Critical/severe vs mild/moderate |
| Bravi et al,<sup>37</sup> 2020 | Italy | Case-control | 543<sup>a</sup> | Patients with COVID-19 and hypertension | NR | NR | Hypertension (100%) | 83% | COVID-19 severity | NR | Severe or very severe/lethal vs mild |
| Mehta et al,<sup>40</sup> 2020 | USA | Cohort | 1735 | Patients with COVID-19 | NR | 57% | Hypertension (93%), diabetes (46%), CAD (22%), HF (17%) | 12% | Hospitalization, ICU admission, mechanical ventilation | NR | NR |
| Yuchen et al,<sup>47</sup> 2020 | China | Cohort | 71<sup>a</sup> | Patients with COVID-19, hypertension, and diabetes | 67 (61-76)<sup>c</sup> | NR | Hypertension (100%), diabetes (100%) | 45% | All-cause mortality, creatinine, procalcitonin, CRP, | In-hospital death | NR |

|  |  |  |  |  |  |  |  |  | IL-6, D-dimer |  |  |
| --- | --- | --- | --- | --- | --- | --- | --- | --- | --- | --- | --- |
| Rhee et al,<sup>38</sup> 2020 | Korea | Cohort | 832 | Patients with COVID-19 and diabetes | NR | 53% | Hypertension (68%), diabetes (100%), CVD (27%), CKD (19%) | 39% | COVID-19 severity | NR | Intensive care or death vs mild |
| Kim et al,<sup>43</sup> 2020 | USA | Cohort | 2491 | Hospitalized patients with COVID-19 | 62 (50-75)<sup>c</sup> | 53% | Hypertension (57%), diabetes (33%), CAD (14%), HF (11%), CKD (16%) | 30% | All-cause mortality, ICU admission | In-hospital death | NR |
| Khera et al,<sup>34</sup> 2020 | USA | Cohort | 10196 | Adult patients (≥18 years) with COVID-19 and hypertension | NR | 54% | Hypertension (100%), diabetes (48%), CAD (5%), HF (27%), CKD (27%) | 59% | All-cause mortality, hospitalization | In-hospital death | NR |
| Jung et al,<sup>33</sup> 2020 | Korea | Cohort | 5179 | Adult patients (≥18 years) with COVID-19 | 44.6 ± 18<sup>b</sup> | 44% | Hypertension (22%), diabetes (17%), CAD (1%), HF (4%), CKD (5%) | 15% | All-cause mortality, mechanical ventilation | In-hospital death | NR |
| Huang et al,<sup>28</sup> 2020 | China | Cohort | 50 | Hospitalized patients with COVID-19 and hypertension | 61.7 ± 12.9<sup>b</sup> | 54% | Hypertension (100%), diabetes (8%), CAD (2%) | 40% | All-cause mortality, COVID-19 severity, mechanical ventilation, troponin, creatinine, procalcitonin in, CRP,<br>D-dimer | In-hospital death | Critical/severe vs mild/moderate |
| De<br>Spiegele<br>er et al,<sup>23</sup><br>2020 | Belgiu<br>m | Cohort | 154 | Residents at two<br>elderly care homes<br>with COVID-19 | 86 ±<br>7<sup>b</sup> | 33% | Hypertension<br>(25%),<br>diabetes<br>(18%) | 20% | COVID-19<br>severity | NR | Long-stay<br>hospital<br>admission<br>or death<br>vs none |
| Baker et<br>al,<sup>82</sup> 2020 | UK | Cohort | 311 | Hospitalized patients<br>with COVID-19 | 75<br>(60-83)<sup>c</sup> | 55% | Hypertension<br>(42%),<br>diabetes<br>(27%), CAD<br>(21%), HF<br>(14%), CKD<br>(24%) | 25% | All-cause<br>mortality | 28-day<br>death | NR |
| Yang et<br>al,<sup>31</sup> 2020 | China | Cohort | 126<sup>a</sup> | Patients with COVID-19 and hypertension | 66<br>(61-73)<sup>c</sup> | 49% | Hypertension<br>(100%),<br>diabetes<br>(30%), CVD<br>(18%) | 34% | All-cause<br>mortality,<br>COVID-19<br>severity,<br>hospital<br>discharge,<br>length of<br>hospital<br>stay,<br>troponin,<br>creatinine,<br>procalciton<br>in, CRP,<br>IL-6, D-dimer | In-hospital<br>death | Critical/se<br>vere vs<br>mild/mode<br>rate |
| Rentsch<br>et al,<sup>19</sup><br>2020 | USA | Cohort | 585 | Patients with<br>laboratory results<br>consistent with<br>SARS-CoV-2 or<br>COVID-19 | 66.1<br>(60.4-71)<sup>c</sup> | 95% | Hypertension<br>(72%),<br>diabetes<br>(44%) | 44% | All-cause<br>mortality,<br>hospitaliza<br>tion, ICU<br>admission | In-hospital<br>death | NR |
| Bean et<br>al,<sup>52</sup> 2020 | UK | Cohort | 205 | Patients with<br>symptoms that<br>required<br>hospitalization with<br>COVID-19 | 63 ±<br>20<sup>b</sup> | 52% | Hypertension<br>(51%),<br>diabetes<br>(30%), CAD<br>(15%) | 23% | COVID-19<br>severity | NR | In-hospital<br>death or<br>required<br>critical<br>care support vs none |
| Feng et al,<sup>24</sup> 2020 | China | Cohort | 65<sup>a</sup> | Consecutive adult patients with COVID-19 | 47 (36-58)<sup>c</sup> | 50% | Hypertension (100%), diabetes (31%), CVD (8%) | 25% | COVID-19 severity, creatinine | NR | Critical/severe vs mild/moderate |
| Zhang et al,<sup>21</sup> 2020 | China | Cohort | 1128 | Patients (18-74 years) with COVID-19 and hypertension | 64 (56-69)<sup>c</sup> | 53% | Hypertension (100%), diabetes (21%), CAD (12%) | 17% | All-cause mortality, mechanical ventilation | 28-day all-cause mortality | NR |
| Giorgi et al,<sup>32</sup> 2020 | Italy | Cohort | 2653 | Symptomatic patients with COVID-19 | 63.2<sup>b</sup> | 50% | Hypertension (18%), diabetes (12%), CAD (7%), HF (6%) | 31% | All-cause mortality, hospitalization | Death | NR |
| Benelli et al,<sup>42</sup> 2020 | Italy | Cohort | 411 | Consecutive patients with COVID-19 | 66.8 ± 16.4<sup>b</sup> | 87% | Hypertension (47%), diabetes (16%), CVD (23%) | 27% | All-cause mortality, ICU admission | In-hospital death | NR |
| Lee et al,<sup>35</sup> 2020 | Korea | Cohort | 8266 | Hospitalized patients with COVID-19 | 44.4 ± 19.1<sup>b</sup> | 38% | Hypertension (19%), diabetes (17%), CAD (6%), HF (1%) | 12% | All-cause mortality | 60-day death | NR |

Abbreviations: ACEIs, angiotensin-converting enzyme inhibitors; ARBs, angiotensin receptor blockers; COVID-19, coronavirus disease 2019; CAD, coronary artery disease; HF, heart failure; CVD, cardiovascular disease; CKD, chronic kidney disease; NR, not reported; CRP, c-reactive protein; IL-6, interleukin-6; ICU, intensive care unit; USA, United States of America; UK, United Kingdom.
<sup>a</sup>Subgroup from original population with data on ACEIs/ARBs use
<sup>b</sup>Mean ± SD
<sup>c</sup>Median (IQR)

Our search in clinicaltrials.gov identified 16 registered RCTs (eTable 3), of which six evaluate the impact of continuation or discontinuation of ACEIs and ARBs on COVID-19 outcomes and four placebo-controlled trials assess the efficacy of ARB (losartan and valsartan) and ACEI (ramipril) in COVID-19 patients who are not previously taking a RAS inhibitor.

### Risk of bias assessment

Almost all cross-sectional studies had moderate risk of bias, all case-control studies had low risk of bias, and 9 of 17 cohort studies had low risk of bias (eTables 4, 5, and 6). None of studies was scored as high risk of bias.

### All-cause mortality

In 22 studies (11 cross-sectional and 11 cohorts, n=23059), the use of ACEIs or ARBs was not associated with higher odds of all-cause mortality (OR 1.11, 95%CI 0.77-1.60, p=0.56) and heterogeneity was high among studies (Figure 2). In the subgroup analysis by study design, ACEIs or ARBs use were not associated with all-cause mortality in 11 cross-sectional studies (OR 0.86, 95%CI 0.65-1.15, p=0.32) and 11 cohort studies (OR 1.30, 95%CI 0.71-2.38, p=0.40) (Figure 2). The funnel plot did not show asymmetry and the Egger’s test was not significant (p=0.39) (eFigure 1). ACEI use was not associated with all-cause mortality (OR 1.18, 95%CI 0.83-1.66, p=0.35) (eFigure 2), but ARB use was associated with increased odds of all-cause mortality (OR 1.79, 95%CI 1.07-3.00, p=0.03) (eFigure 3).

Six studies^21,32,36^ reported adjusted effect measures of association between ACEIs/ARBs use and all-cause mortality (eTable 7). The pooled estimate of three studies^21,35,36^ with similar adjusted variables (age, sex, and cardiovascular comorbidities) found that ACEIs or ARBs use was not associated with all-cause mortality (aHR 0.83, 95%CI 0.49-1.38, p=0.47) (eFigure 4). Furthermore, the pooled estimate of ACEI studies^32,34,35^ (aHR 0.97, 95%CI 0.83-1.13, p=0.67) and ARB studies^32,34,35^ (aHR 1.14, 95%CI 0.98-1.34, p=0.08) showed no association either (eFigure 5 and 6).

**Figure 2.**
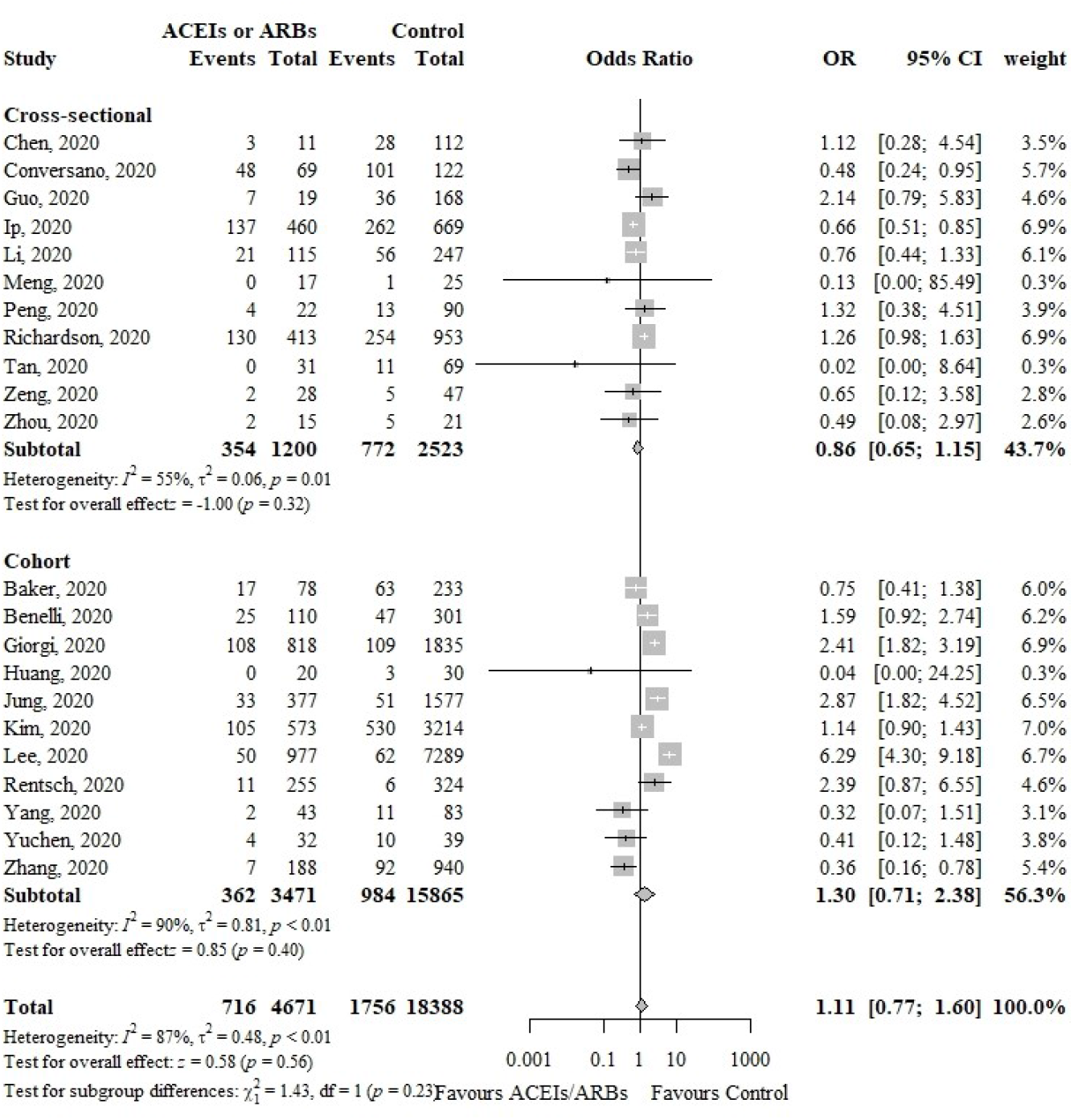
Forest plot showing the association between ACEIs or ARBs use and all-cause mortality in COVID-19 patients.

### Secondary Outcomes

#### CO VID-19 severity

In 18 studies (11 cross-sectional, two case-control, and five cohorts, n=11870), the use of ACEIs or ARBs was not associated with COVID-19 severity (OR 0.79, 95%CI 0.59-1.07, p=0.13) and showed high heterogeneity among studies (Figure 3). The funnel plot showed asymmetry, suggesting publication bias which was confirmed by the Egger’s test (p<0.01) (eFigure 7). Subgroup analysis by study design showed that ACEIs or ARBs use was only associated with lower COVID-19 severity in five cohort studies (OR 0.61, 95%CI 0.39-0.95, p=0.03) (Figure 3). In contrast, the use of ACEI (OR 1.10, 95%CI 0.55-2.18, p=0.79) and ARB (OR 1.00, 95%CI 0.77-1.29, p=0.98) separately were not associated with COVID-19 severity (eFigure 8 and 9).

**Figure 3.**
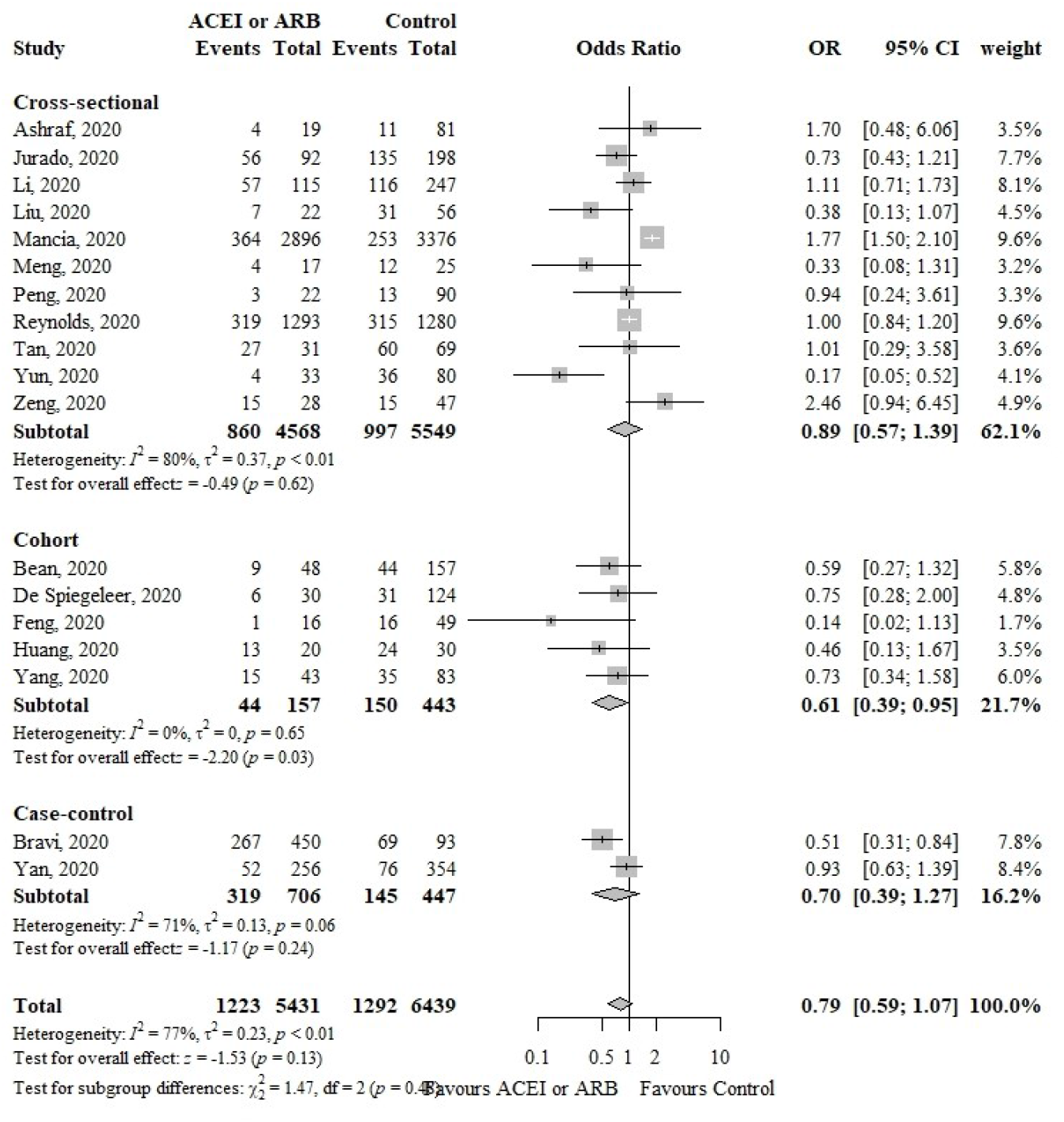
Forest plot showing the association between ACEIs or ARBs use and COVID-19 severity.

The pooled adjusted estimate of four studies^23,24,37,38^ showed that the ACEIs or ARBs use (aOR 0.56, 95%CI 0.37-0.87, p<0.01) was independently associated with lower COVID-19 severity (eFigure 10). However, adjusted estimates of ACEI (aOR 0.66, 95%CI 0.37-1.18, p=0.15) and ARB (aOR 0.97, 95%CI 0.79-1.20, p=0.81) use separately were not associated with COVID-19 severity (eFigure 11 and 12).

#### Hospital discharge

In three studies^29,31,39^ (two cross-sectional and one cohort, n=301), the use of ACEIs or ARBs was not associated with hospital discharge (OR 2.27, 95%CI 0.96-5.35, p=0.06) (eFigure 13).

#### Hospitalization

In four studies^19,32,39,40^ (one cross-sectional and three cohorts, n=5048), the use of ACEIs or ARBs was not associated with hospitalization (OR 1.83, 95%CI 0.95-3.52, p=0.07) (eFigure 14). Likewise, the use of ACEI (OR 1.63, 95%CI 0.94-2.83, p=0.08) and ARB (OR 1.48, 95%CI 0.95-2.31, p=0.08) were not associated with hospitalization (eFigure 15 and 16).

#### ICU admission

In six studies^19,40-44^ (two cross-sectional and four cohorts, n=8884), the use of ACEIs or ARBs was associated with increased odds of ICU admission (OR 1.45, 95%CI 1.17-1.80, p<0.01) (eFigure 17). In contrast, the use of ACEI (OR 1.16, 95%CI 0.72-1.86, p=0.53) and ARB (OR 1.26, 95%CI 0.87-1.83, p=0.23) by separate were not associated with ICU admission (eFigure 18 and 19).

#### Mechanical Ventilation

In seven studies^21,28,29,33,40,44,45^ (three cross-sectional and four cohorts, n=6533), the use of ACEIs or ARBs was not associated with mechanical ventilation (OR 1.39, 95%CI 0.99-1.94, p=0.06) (eFigure 20).

#### Length of hospital stay

In five studies^26,29,31,39,46^ (four cross-sectional and one cohort, n=699), the use of ACEIs or ARBs was not associated with length of hospital stay (MD −0.96 days, 95%CI −2.50 to 0.57, p=0.22) (eFigure 21).

#### Troponin level

In four studies^26-28,31^ (two cross-sectional and two cohorts, n=580), the use of ACEIs or ARBs was not associated with troponin level (MD −0.01 μg/L, 95%CI −0.04 to 0.02, p=0.37) (eFigure 22).

#### Creatinine level

In six studies^24,26-28,31,47^ (two cross-sectional and four cohorts, n=716), the use of ACEIs or ARBs was not associated with creatinine level (MD −0.58 μmol/L, 95%CI −8.72 to 7.56, p=0.89) (eFigure 23).

#### Procalcitonin level

In five studies^26-28,31,47^ (two cross-sectional and three cohorts, n=651), the use of ACEIs or ARBs was not associated with procalcitonin level (MD −0.02 ng/mL, 95%CI −0.05 to 0.01, p=0.21) (eFigure 24).

#### CRP level

In five studies^26,28,29,31,47^ (two cross-sectional and three cohorts, n=709), the use of ACEIs or ARBs was not associated with CRP level (MD −6.39 mg/L, 95%CI −16.19 to 3.41, p=0.20) (eFigure 25).

#### IL-6 level

In four studies^26,27,31,47^ (two cross-sectional and two cohorts, n=601), the use of ACEIs or ARBs was not associated with IL-6 level (MD −4.41 pg/mL, 95%CI −13.24 to 4.42, p=0.33) (eFigure 26).

#### D-dimer level

In six studies^26,29,31,47^ (two cross-sectional and one cohort, n=751), the use of ACEIs or ARBs was not associated with D-dimer level (MD −0.91 nmol/L, 95%CI −2.77 to 0.94, p=0.33) (eFigure 27).

### Sensitivity analyses

The results of the sensitivity analyses are reported in the eTable 8. Overall, the results showed that ACEIs or ARBs use was independently with lower COVID-19 severity, ARB use was independently associated with higher mortality, and ACEIs or ARBs use was associated with higher ICU admission.

## Discussion

We found that the use of ACEIs or ARBs was not significantly associated with all-cause mortality in COVID-19 patients, and when analyzed by study design or when using adjusted effects. In contrast, ACEIs or ARBs use was independently associated with lower COVID-19 severity. Although ACEIs or ARBs use was associated with an increased odds of ICU admission, this effect disappeared when ACEIs and ARBs were analyzed individually. No significant associations were found between ACEIs or ARBs use and other clinical outcomes or biomarkers. Risk of bias was low or moderate across studies.

It has been proposed that RAS play a crucial role in the pathogenesis of infection by SARS-CoV-2, since it uses the ACE2 receptor to enter into cells, with the subsequent downregulation of this surface protein.^5^ The reduction of ACE2 expression in infected cells can lead to a tissue and systemic RAS imbalance with a predominance of the dangerous ACE/Ang II/AT_1_R axis.^5^ This phenomenon can be particularly harmful in the elderly population since they have already a lower level of ACE2 expression compared to young people.^48^ This could partly explain the higher mortality observed in older patients with COVID-19.^49^ Recent evidence from a Chinese cohort of 12 COVID-19 patients showed that circulating Ang II levels were markedly elevated compared to healthy controls and linearly associated with viral load and lung injury.^50^ Moreover, in an animal experiment of acute lung injury induced by acid, the SARS-CoV spike protein enhances the pulmonary Ang II levels and lung injury severity.^51^ Altogether, these data suggest that SARS-CoV-2 can mediate the damage to lungs and possibly to other organs through the absence of degradation of Ang II. Therefore, RAS modulators such as ACEIs and ARBs can be used as potential therapeutic agents in COVID-19 patients. This is currently under investigation in several ongoing clinical trials.

In general, we found that the use of ACEIs or ARBs had a neutral effect on all-cause mortality and other clinical outcomes in COVID-19 patients. Two studies^21,52^ with larger samples and adjustment for confounders reported a significant reduction of all-cause mortality and severity in these patients. Nowadays, there is controversy regarding the use of ACEIs and ARBs in patients with COVID-19 and hypertension. Initially it was suggested that the use of these drugs could increase ACE2 expression; however, there is conflicting evidence about the effect of ACEIs and ARBs on ACE2 tissue expression in animal models.^53^ Besides, studies in humans showed no effect of ACEIs and ARBs administration on ACE2 protein levels in urine and plasma.^54,55^ Furthermore, a recent Mendelian randomization study revealed a lack of association between genetically proxy ACE inhibition and lung ACE2 expression or circulating ACE2 levels.^56^ Likewise, in a study on human myocardial samples, there was no significant difference in ACE2 expression in tissue samples with and without exposure to ACEI.^57^ Overall, these findings suggest that ACEIs and ARBs are unlikely to raise ACE2 in humans. Thus, it seems reasonable that ACEIs and ARBs could exert its effect on COVID-19 mainly through inhibition of the ACE/Ang II/AT_1_R axis.

The lung is the target organ in COVID-19; however, other organs may potentially be involved. A recent study reported that acute cardiac injury, manifested as elevated troponin levels, was present in 20% of COVID-19 patients and was independently associated with worse outcomes.^58^ Although the pathophysiological basis of this finding is not fully understood, it has been proposed that SARS-CoV-2 can cause cardiac injury through several mechanisms: direct viral damage, systemic inflammatory response, microangiopathy, and myocardial infarction.^59^ Similarly, acute kidney injury was observed in up to 27% of COVID-19 patients,^60^ this is probably related to alterations in renal microvasculature, kidney cell viral infection, and systemic inflammation.^61^ Dysregulation of the immune system is key in the pathogenesis of COVID-19 leading, in some cases, to an overproduction of pro-inflammatory cytokines (interleukin-6, interleukin-1β, and tumor necrosis factor-alpha) resulting in what has been called a cytokine storm.^62^ Likewise, the procoagulant-anticoagulant balance has been found to be impaired in COVID-19, leading to formation of microthrombi and marked elevation of D-dimer.^63^

A recent meta-analysis found that elevation of troponin, creatinine, D-dimer, and procalcitonin were significantly associated with a higher risk of critical disease or mortality in COVID-19 patients.^64^ The RAS imbalance and the loss of ACE2 expression observed in COVID-19, with the subsequent reduction of Ang 1-7 and elevation of Ang II levels, can contribute to the tissue and systemic damage caused by SARS-CoV-2. Thus, considering that RAS inhibitors are capable of regulating both tissue and systemic RAS, it has been suggested that could have a beneficial effect in COVID-19. However, our study did not find a significant association of ACEIs or ARBs use on troponin, inflammatory markers (procalcitonin, CRP, and IL-6), creatinine, and D-dimer levels in COVID-19 patients. Further research is needed to clarify the potential therapeutic role of RAS inhibitors on multiorgan dysfunction associated with COVID-19.

We excluded two large observational studies by Mehra et al.^65^ (n=8910) and by Mehra et al.^66^ (n=96032) because of several concerns of the quality of their registry data in open letters by researchers worldwide^67^ and acknowledged by the two journals in expressions of concern.^68,69^

There are three previous systematic reviews examining the effects of ACEI/ARB use on COVID-19 patients. Zhang et al.^70^ found that ACEI/ARB exposure was not associated with a higher risk of severe infection or mortality. However, only 12 studies and unadjusted estimates were combined. Guo et al.^71^ showed that ACEI/ARB use was associated with lower mortality in COVID-19 patients although only included six studies were included. Mackey et al.^72^ only conducted a narrative synthesis of 14 studies, concluding that there is no evidence of association between ACEI/ARB use with more severe COVID-19 disease. Compared to these reviews, our study included 40 studies and evaluated 13 outcomes. Additionally, to our knowledge, our review is the first that pooled adjusted effect estimates for mortality and COVID-19 severity.

Our study has some limitations. First, given most of the studies did not use adjusted effects, there is an increased risk of bias in their pooled effect measures. Thus, these results should be considered with caution. However, we also reported meta-analyses of adjusted estimates of a few available studies. Second, the majority of the included studies were of cross-sectional design, thus causality cannot be concluded due to the methodological limitations of this design. Third, heterogeneity was high in most of the evaluated outcomes. Possible reasons for heterogeneity include sample size, differences in outcome definitions, heterogeneous population, among others. Fourth, given that discontinuation of ACEIs or ARBs during hospitalization was not reported consistently across studies, this could influence the significance of pooled estimates. Finally, we could not adequately evaluate the effects of ACEIs and ARBs by separate, since were mainly reported as aggregate due to scarcity of studies.

## Conclusions

In conclusion, the use of ACEIs or ARBs was not associated with higher all-cause mortality in COVID-19 patients, based on the meta-analysis of cross-sectional and cohort studies and also using adjusted effects. ACEI or ARB use was independently associated with lower COVID-19 severity. Also, there was no evidence of association between ACEIs or ARBs use and nearly all secondary clinical outcomes and biomarkers. Although these results are not conclusive, our review supports current international guidelines to continue the use of RAS inhibitors in COVID-19 patients with hypertension. More studies are needed to determine the potential beneficial effect of ACEIs and ARBs in patients with COVID-19.

## Data Availability

NA

## Author Contributions

Diaz-Arocutipa had full access to all of the data in the study and takes responsibility for the integrity of the data and the accuracy of the data analysis. *Study concept and design:* Diaz-Arocutipa. *Acquisition, analysis, or interpretation of data:* Diaz-Arocutipa, Saucedo-Chinchay, Hernandez. *Drafting of the manuscript:* Diaz-Arocutipa. *Critical revision of the manuscript for important intellectual content:* Diaz-Arocutipa, Saucedo-Chinchay, Hernandez. *Statistical analysis:* Diaz-Arocutipa, Hernandez. *Administrative, technical, or material support:* Diaz-Arocutipa. *Study supervision:* Diaz-Arocutipa, Hernandez.

